# Association Between Area Deprivation and Dental Provider Density in California: A Cross-Sectional Ecological Study

**DOI:** 10.64898/2026.07.04.26357261

**Authors:** Anna-Lena Asiedu, Collins Gaba

**Affiliations:** University of California, San Francisco School of Dentistry, San Francisco, CA, USA; Center for Innovation in Social Work and Health, Boston University School of Social Work, Boston, MA, USA

**Keywords:** Area Deprivation Index, dental provider density, dental workforce, oral health disparities, access to dental care, dental deserts, California

## Abstract

**Background:** Neighborhood socioeconomic disadvantage may contribute to inequities in access to dental care by influencing the geographic distribution of providers. The Area Deprivation Index (ADI) is a validated measure of neighborhood deprivation, but its association with dental workforce availability has not been examined statewide in California. This study evaluated the relationship between neighborhood deprivation and dental provider density across California ZIP Code Tabulation Areas (ZCTAs).

**Methods:** We conducted a cross-sectional ecological study of California ZCTAs using publicly available data from the National Plan and Provider Enumeration System (April 2026), the Neighborhood Atlas 2023 ADI, and 2024 U.S. Census population estimates. Active dental providers were linked to ZCTAs and provider density was calculated per 10,000 residents. ADI was aggregated to the ZCTA level using the median ADI national percentile. Negative binomial regression was used to assess the association between ADI and dental provider density, with population included as an offset. Secondary analyses examined California-specific ADI quartiles, dental deserts, and specialist versus general dentist availability.

**Results:** The final analytic sample included 1,426 California ZCTAs representing 39,016,384 residents and 37,945 active dental providers. Greater neighborhood deprivation was significantly associated with lower dental provider density. Each one-percentile increase in ADI corresponded to a 1.8% reduction in provider density (incidence rate ratio [RR] 0.9823, 95% confidence interval [CI] 0.9799–0.9847; p < 0.001). Compared with the least deprived quartile, the most deprived quartile had 61% fewer dental providers (RR 0.39, 95% CI 0.34–0.45; p < 0.001). Overall, 15.9% of ZCTAs contained no active dental providers, increasing from 6.8% in the least deprived quartile to 31.1% in the most deprived quartile. Specialist availability demonstrated an even steeper deprivation gradient, with specialist density declining by 86% between the least and most deprived quartiles.

**Conclusions:** Neighborhood deprivation was strongly associated with reduced dental provider availability across California. Highly deprived communities had substantially lower provider density, were more likely to be dental deserts, and experienced particularly pronounced shortages of dental specialists. ADI may be a useful tool for identifying communities where workforce shortages and socioeconomic disadvantage intersect and for informing equitable workforce planning and policy interventions.

**Trial registration:** Not applicable. This study used publicly available, de-identified data and did not involve human participants.

## Introduction

Access to dental care depends not only on insurance coverage and patient demand but also on the geographic distribution of the dental workforce (1,2). Although California has the largest dental workforce of any U.S. state, dental providers are unevenly distributed, leaving some communities with substantially lower provider density than others (3,4,5). This geographic variation suggests that where people live influences access to dental care (2). However, contemporary neighborhood-level evidence describing how dental provider availability varies across neighborhoods with different levels of socioeconomic disadvantage in California remains limited. Characterizing these patterns may help identify communities with the greatest unmet need and inform workforce planning, resource allocation, and oral health policy.

The importance of workforce distribution extends beyond California. Nationally, an estimated 24.7 million people reside in dental care deficit areas, and substantial urban-rural disparities in dental provider availability persist (2). Although workforce projections suggest that the overall national supply of dentists may be adequate in the coming decades, aggregate workforce numbers obscure important geographic maldistribution (2,3). Communities with the greatest burden of oral disease often have the fewest available providers, creating a persistent cycle of unmet need (1,2,6).

Understanding how dental provider distribution aligns with neighborhood socioeconomic deprivation is important because disparities extend beyond the traditional rural–urban divide (1,7). Individuals living in deprived communities often face multiple barriers to obtaining dental care, including transportation limitations, reduced provider availability, insurance network shortages, and lower provider participation in public insurance programs (1,6,7). Neighborhood deprivation captures broader contextual factors beyond individual income, encompassing education, employment, housing quality, and other social conditions that shape opportunities to access healthcare throughout the life course (8–10). Evaluating provider distribution in relation to neighborhood deprivation may therefore provide a more comprehensive understanding of dental workforce inequities than geographic location alone.

The Area Deprivation Index (ADI) is a validated composite measure of neighborhood socioeconomic disadvantage derived from 17 census-based indicators of income, education, employment, and housing quality (9–12). ADI has become an important tool for examining how neighborhood socioeconomic disadvantage influences healthcare access and health outcomes (9–12). Previous studies have demonstrated associations between neighborhood deprivation and poorer oral health outcomes and have identified geographic disparities in dental workforce availability (2,13,14,15). However, most investigations have relied on county-level analyses, social vulnerability indices rather than the Area Deprivation Index (ADI), commercial provider databases, or outcomes focused on dental care utilization rather than provider availability (2,14). As a result, important neighborhood-level variation in dental workforce distribution may remain obscured (10,16).

In addition to having the nation’s largest dental workforce, California administers the largest Medicaid dental program in the United States (17). However, insurance coverage alone does not guarantee access when provider participation remains uneven across communities (1,17). Moreover, previous California studies evaluating dental workforce distribution were conducted more than two decades ago, before the availability of the Area Deprivation Index (ADI) and the National Plan and Provider Enumeration System (NPPES) (4,18).

Using statewide NPPES provider data, Neighborhood Atlas ADI data, and U.S. Census population estimates linked at the ZCTA level, this study examined the association between neighborhood deprivation and dental provider density across California. By characterizing dental workforce distribution at the neighborhood level using publicly available, reproducible datasets, this study provides contemporary evidence to support geographically targeted workforce planning, resource allocation, and policy efforts to improve equitable access to dental care.

## Methods

### Study Design

This study is a cross-sectional, ecological analysis of the association between neighborhood-level area deprivation and dental provider density across the state of California. The unit of analysis was the 5-digit ZCTA, the United States Census Bureau’s geographic approximation of a 5-digit ZIP code. The study used only publicly available, de-identified, aggregated data and was therefore not human-subjects research; institutional review board approval was not required. Reporting followed the Strengthening the Reporting of Observational Studies in Epidemiology (STROBE) guidelines (19).

### Data Sources

#### Dental Providers

We identified active dental providers in California from the National Plan and Provider Enumeration System (NPPES) database, April 2026 release. NPPES is the federal registry maintained by the Centers for Medicare & Medicaid Services that assigns a unique 10-digit National Provider Identifier (NPI) to all U.S. health care providers and is updated continuously as providers report changes to practice location, credentials, and licensure status (20). We retained individual providers with an active NPI, a California practice address, and at least one of fourteen dental taxonomy codes from the National Uniform Claim Committee Health Care Provider Taxonomy Code Set version 26.0, covering general dentistry and all twelve American Dental Association–recognized specialties (see Supplementary Table S1 for the full taxonomy list and per-taxonomy counts in the final analytic sample) (21). Each provider was classified by their first matching dental taxonomy code and de-duplicated by NPI.

#### Area Deprivation Index

Neighborhood deprivation was measured using the 2023 Area Deprivation Index (ADI) 9-Digit ZIP Code File, version 4.0.1 from the Neighborhood Atlas at the University of Wisconsin–Madison. The ADI is a validated census-derived composite measure constructed from 17 indicators of income, education, employment, and housing quality at the Census block-group level, reported as a national percentile (1–100) and a state-specific decile (1–10), with higher values indicating greater deprivation (9,12,22). Records with Neighborhood Atlas suppression codes (GQ, PH, GQ-PH, QDI, P, U) were excluded from ADI computation. ADI is provided at two units: the census block group and the 9-digit ZIP+4. NPPES records dental provider location by ZIP code, and Census population estimates are released at the ZCTA level. The 9-digit ZIP+4 file is the Neighborhood Atlas’s recommended source for ZIP-based research, constructed by the Atlas team as a direct crosswalk to the block-group–level ADI (12, 22). To link these variables, the ADI was aggregated from the 9-digit ZIP+4 level to the 5-digit ZCTA by taking the median of all non-suppressed block-group scores within each ZCTA; the median, rather than the mean, was used to reduce the influence of outlying block groups within heterogeneous ZCTAs. The Neighborhood Atlas notes that linking ADI values to geographic units larger than the block group is not a validated approach (22); this trade-off was unavoidable as the three data sources (NPPES, ADI, Census population) could not be linked at any common finer geography.

#### Population

ZCTA-level population estimates for California were obtained from the United States Census Bureau’s 2024 population estimates (23). Population was used to quantify the dental provider density (dentists per 10,000 residents).

### Study Sample

Three data sources were linked at the 5-digit ZCTA level. The 5-digit ZCTA was selected as the analytic unit because U.S. Census population estimates are available only at the ZCTA level, and because 11.8% of providers had only 5-digit ZIP codes recorded in the National Plan and Provider Enumeration System (NPPES), precluding complete linkage at the 9-digit level. Active California dental providers identified in NPPES (n = 39,049) were merged with the U.S. Census Bureau 2024 California ZCTA population file. Providers whose practice ZIP codes were not represented in the Census ZCTA file (n = 220) were excluded because no corresponding population denominator was available. The merged dataset consisted of 38,829 active dental providers across 1,803 California ZCTAs.

ZCTAs unsuitable for population-based dentist density estimation were subsequently excluded if they lacked a meaningful residential population denominator or a valid Area Deprivation Index (ADI) measure. Exclusions were applied for three reasons. First, 40 ZCTAs with a Census population of zero (typically institutional or non-residential ZIPs such as university campuses) were excluded, removing 162 providers. Second, 232 ZCTAs with populations below 500 were excluded, consistent with prior ZCTA level dental access studies that excluded sparsely populated ZCTAs using similar population thresholds (24). This criterion removed 309 providers (0.8% of the merged dataset). Third, 105 additional ZCTAs were excluded because all constituent block groups had suppressed ADI values, preventing construction of a reliable median ADI score; this removed an additional 413 providers.

The final analytic sample consisted of 1,426 California ZCTAs representing 39,016,384 residents and 37,945 active dental providers.

### Outcomes and Exposures

The primary outcome was the count of active dental providers per ZCTA, modeled with log(population) as an offset so that effect estimates are interpretable as rates per resident. The primary exposure was the ZCTA-level median ADI national percentile, modeled both as a continuous variable and as California-specific empirical quartiles. We constructed California-specific ADI quartiles because California ZCTAs are concentrated at the lower end of the national ADI distribution (median = 17). Using fixed national cut points placed 73.3% of California ZCTAs in the least deprived national tertile, limiting contrast for between-group comparisons. We report fixed national tertiles descriptively for comparability with other studies.

### Statistical Analysis

Model selection was guided by distributional diagnostics. The rate outcome was severely right-skewed (skewness = 6.10), and the count outcome was substantially overdispersed (variance-to-mean ratio = 36.87), indicating that ordinary least squares and standard Poisson regression were not optimal modeling approaches. Negative binomial regression was therefore selected as the primary analytic approach. Findings were evaluated through a distributional robustness check, a categorical exposure specification, and two pre-specified sensitivity analyses.

### Primary Analysis

A negative binomial regression model was fitted with the count of dental providers per ZCTA as the outcome, the median ADI national percentile as a continuous exposure, and log(population) included as an offset. Effect estimates are reported as rate ratios (RRs) with 95% confidence intervals; a rate ratio below 1 indicates lower dental provider density at higher levels of neighborhood deprivation.

### Robustness Check (Distributional Family)

To verify that the association did not depend on the choice of model family, we fitted a log-linear ordinary least squares regression model with log(dentists per 10,000 + 0.1) as the outcome. Results were unchanged when alternative constants of 0.01 and 1.0 were used.

### Categorical Specification (Dose-Response)

To characterize the dose-response relationship across the deprivation distribution, the primary negative binomial model was refitted using California-specific empirical ADI quartiles in place of the continuous exposure, with the least-deprived quartile as the reference.

### Sensitivity Analyses

Two pre-specified sensitivity analyses repeated the primary model. First, we excluded Travis Air Force Base (ZIP 94535), the single military installation in the sample with embedded dental services, to assess whether this outlying ZCTA influenced the population-level estimate. Second, we replaced the continuous ADI national percentile with the California-specific ADI state decile to assess whether findings were sensitive to the choice of ADI scaling.

### Provider-Type Sub-Analysis

To characterize workforce availability by provider type, general dentists (taxonomy codes 122300000X and 1223G0001X) and dental specialists (the remaining twelve dental taxonomy codes) were analyzed separately in a descriptive sub-analysis. For each provider type, ZCTA-level density (providers per 10,000 residents) and the prevalence of ZCTAs with no provider of that type were summarized overall and by California-specific empirical ADI quartile; no inferential models stratified by provider type were fit.

### **County-Level** Visualization

For visualization, ZCTA-level results were additionally aggregated to the California county level using the Census Bureau ZCTA-to-county relationship file and mapped as choropleths. County-level results are presented for descriptive purposes only; the ZCTA remained the unit of analysis for all inferential models.

All analyses were conducted in R version 4.3.3. Statistical significance was based on a two-sided *P* < 0.05.

## Results

### Sample Characteristics

The final analytic sample comprised 1,426 California ZCTAs, covering 39,016,384 residents (approximately 99.6% of the 2024 California population) and 37,945 active dental providers identified in NPPES. Across the analytic sample, the mean dental provider density was 9.58 per 10,000 residents (SD 14.56; median 6.36; interquartile range 2.44 to 11.92). Provider density was severely right-skewed (skewness = 6.10; kurtosis = 58.39), and the underlying count of providers per ZCTA was substantially overdispersed (variance-to-mean ratio = 36.87). The distributions of dental provider density and ADI national rank are shown in Figures 1 and 2.

The ADI distribution across California ZCTAs was concentrated toward the less-deprived end of the national scale, with a median ZCTA-level ADI national rank of 17 (mean 23.3; interquartile range 7 to 36) and a corresponding median California-specific state decile of 6.

**Figure 1.**
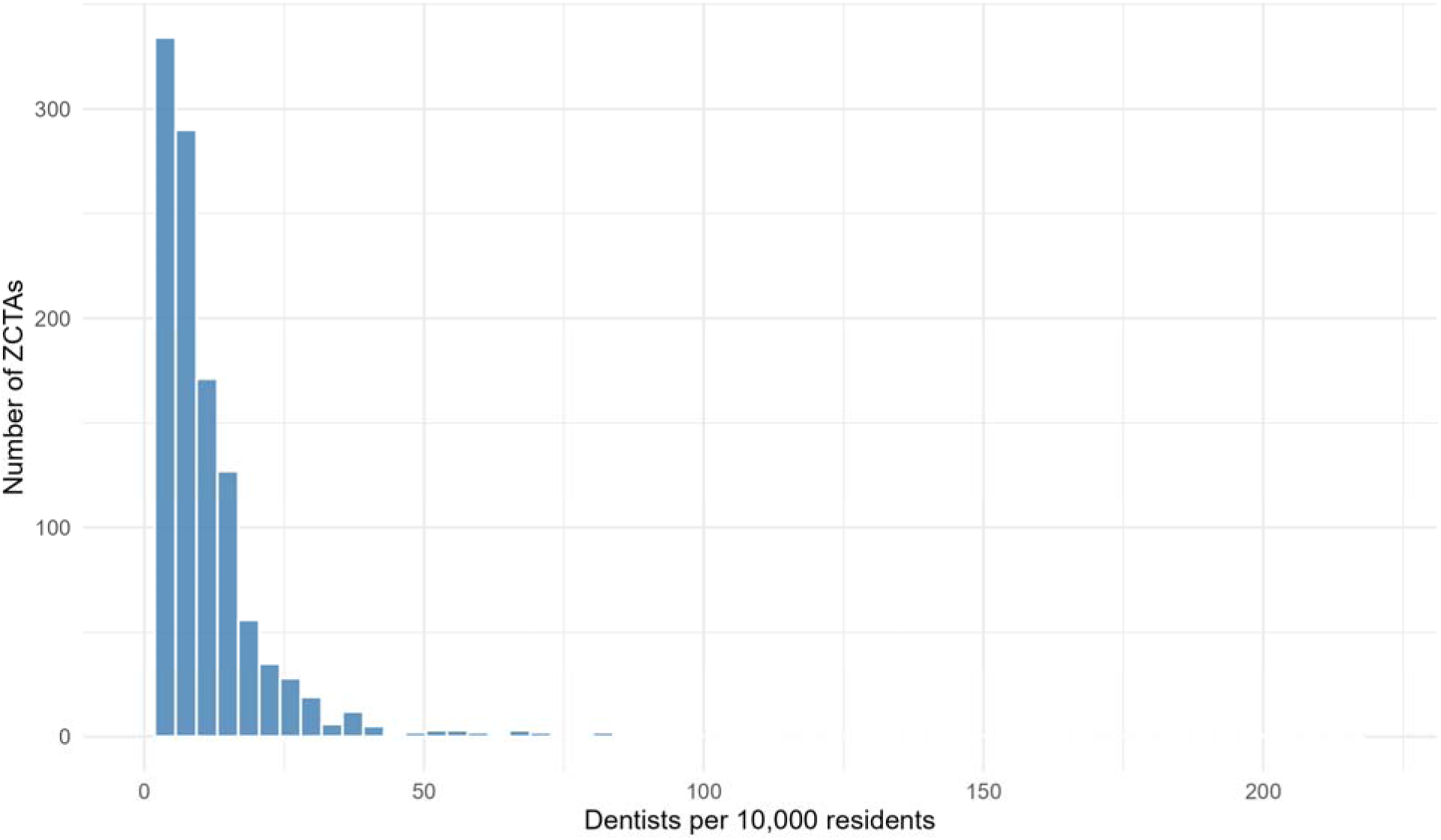
Distribution of dental provider density across California ZCTAs. *Note: Distribution of dental provider density across California ZCTAs in the final analytic sample (n = 1,426)*

**Figure 2.**
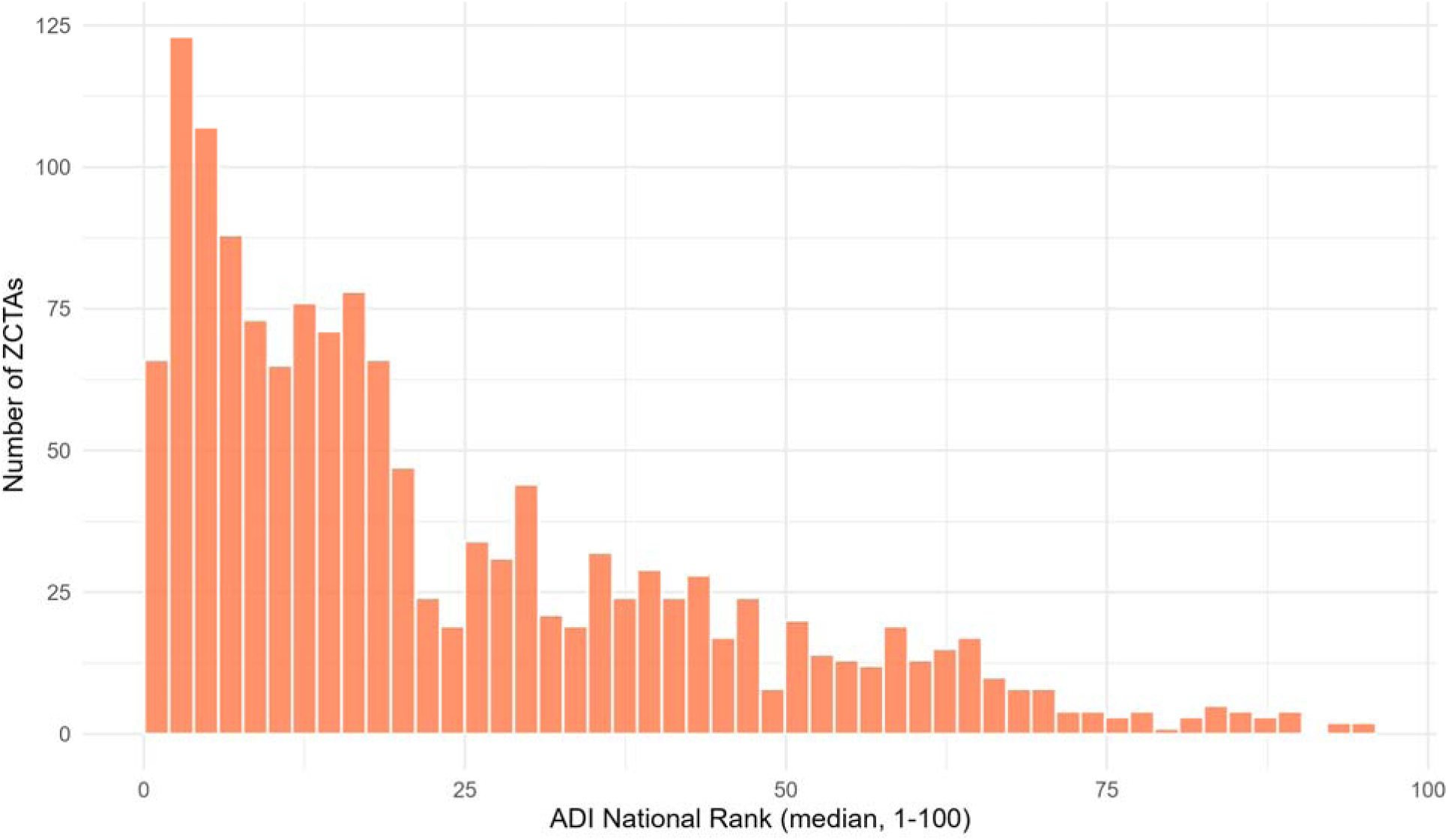
Distribution of ZCTA-level median ADI national rank across California ZCTAs. *Note: Median ADI National Percentile per ZCTA (higher = more deprived)*

### Association Between Area Deprivation and Dental Provider Density

In the primary negative binomial model with the median ADI national percentile as a continuous exposure, each one-percentile increase in ADI was associated with a 1.8% reduction in dental provider density (RR 0.9823; 95% CI 0.9799 to 0.9847; p < 0.001). To illustrate the magnitude of the association, a 25-percentile increase in ADI corresponded to an approximately 36% lower provider density (RR 0.9823^25 ≈ 0.64), and a 50-percentile increase corresponded to an approximately 59% lower density (RR ≈ 0.41). The Locally Estimated Scatterplot Smoothing (LOESS) smoother and linear fit overlaid on the ZCTA-level scatterplot (Figure 3) showed a monotonic negative gradient that closely tracked the linear fit, consistent with the constant multiplicative effect assumed by the negative binomial model.

**Figure 3.**
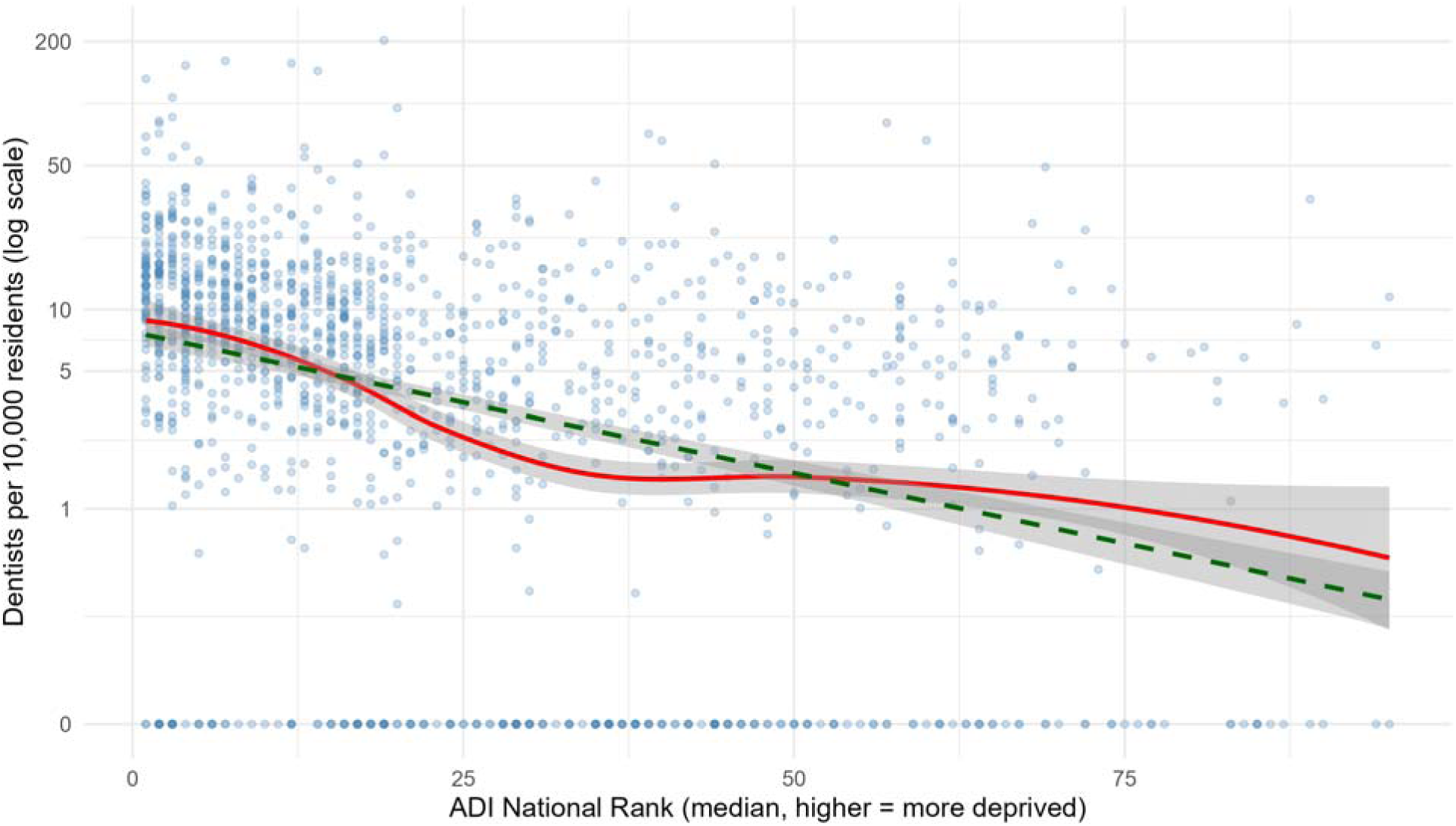
Association between ZCTA-level median ADI national rank and dental provider density. *Note: (LOESS smoother in red; linear fit in green dashed). The y-axis is on a log scale to match the log-link of the negative binomial model; original units are displayed at tick marks. All 1,426 ZCTAs are included; ZCTAs with zero dental providers are plotted using a log(0 + 0.1) constant*.

### Dose-Response Across California-Specific Deprivation Quartiles

When ADI was modeled as California-specific empirical quartiles of the national ADI (Q1: ADI 1–7; Q2: 8–17; Q3: 18–36; Q4: 37–95), a monotonic dose-response gradient emerged. Compared with Q1 (the least deprived), Q2 had 28% fewer dental providers per resident (RR 0.72; 95% CI 0.63 to 0.81; p < 0.001), Q3 had 49% fewer (RR 0.51; 95% CI 0.44 to 0.58; p < 0.001), and Q4 had 61% fewer (RR 0.39; 95% CI 0.34 to 0.45; p < 0.001). Descriptively, mean provider density declined from 14.48 per 10,000 in Q1 to 5.50 per 10,000 in Q4, and median density declined from 10.36 to 2.90 per 10,000 (Figure 4).

**Figure 4.**
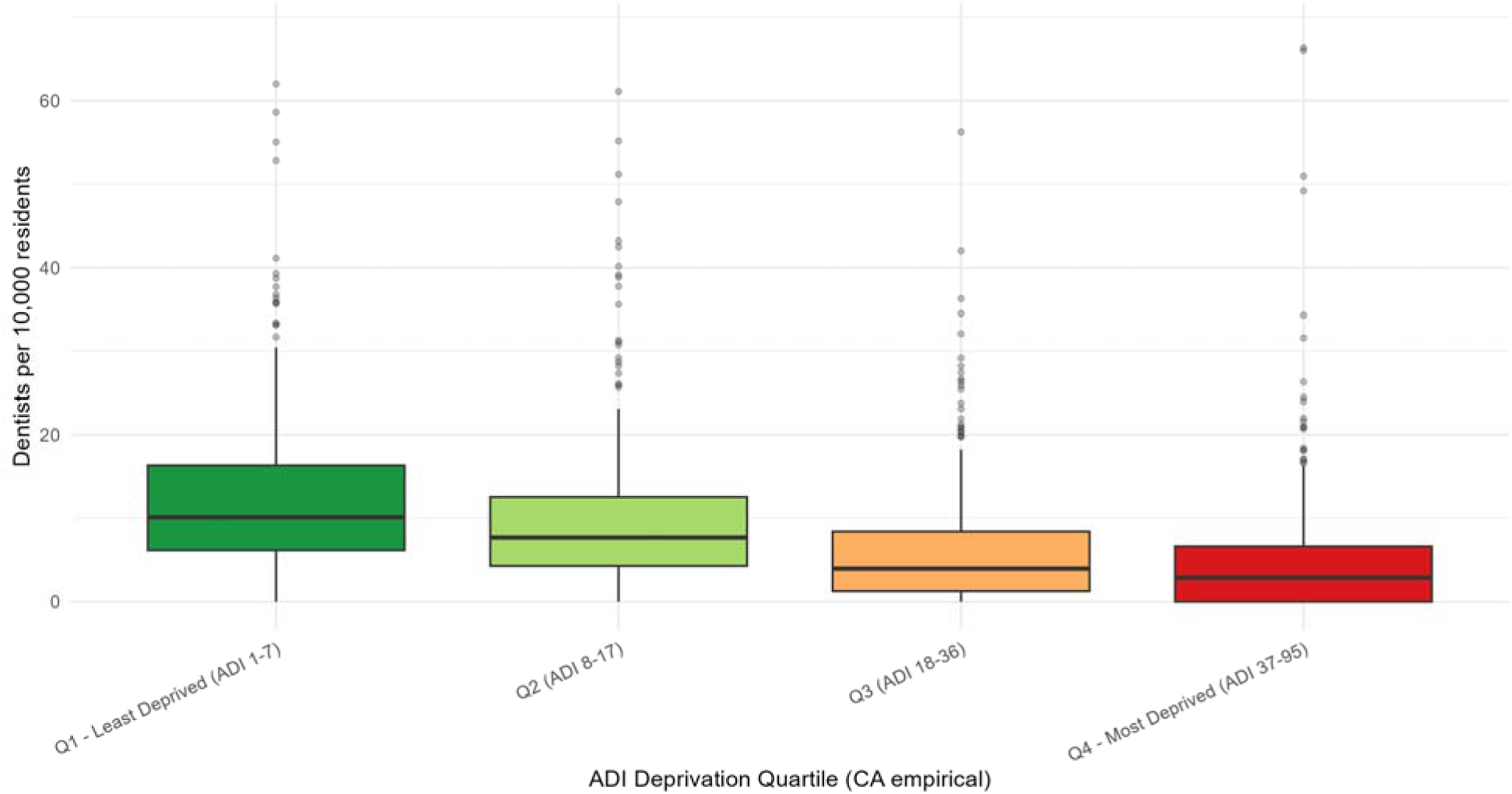
Dental provider density by California-specific empirical ADI deprivation quartile. *Note: Y-axis capped at the 99th percentile*.

### Dental Deserts by Deprivation Quartile

Overall, 227 of 1,426 ZCTAs (15.9%) contained no active dental providers. The prevalence of these dental deserts varied substantially across deprivation quartiles. Among the least-deprived quartile (Q1), 6.8% of ZCTAs had no dental provider; the second quartile was similar at 6.6%. The proportion increased sharply at higher levels of deprivation: 20.5% of Q3 ZCTAs and 31.1% of Q4 ZCTAs contained no dental provider (Figure 5). ZCTAs in the most-deprived California quartile were approximately 4.6 times more likely to be dental deserts than those in the least-deprived quartile.

**Figure 5.**
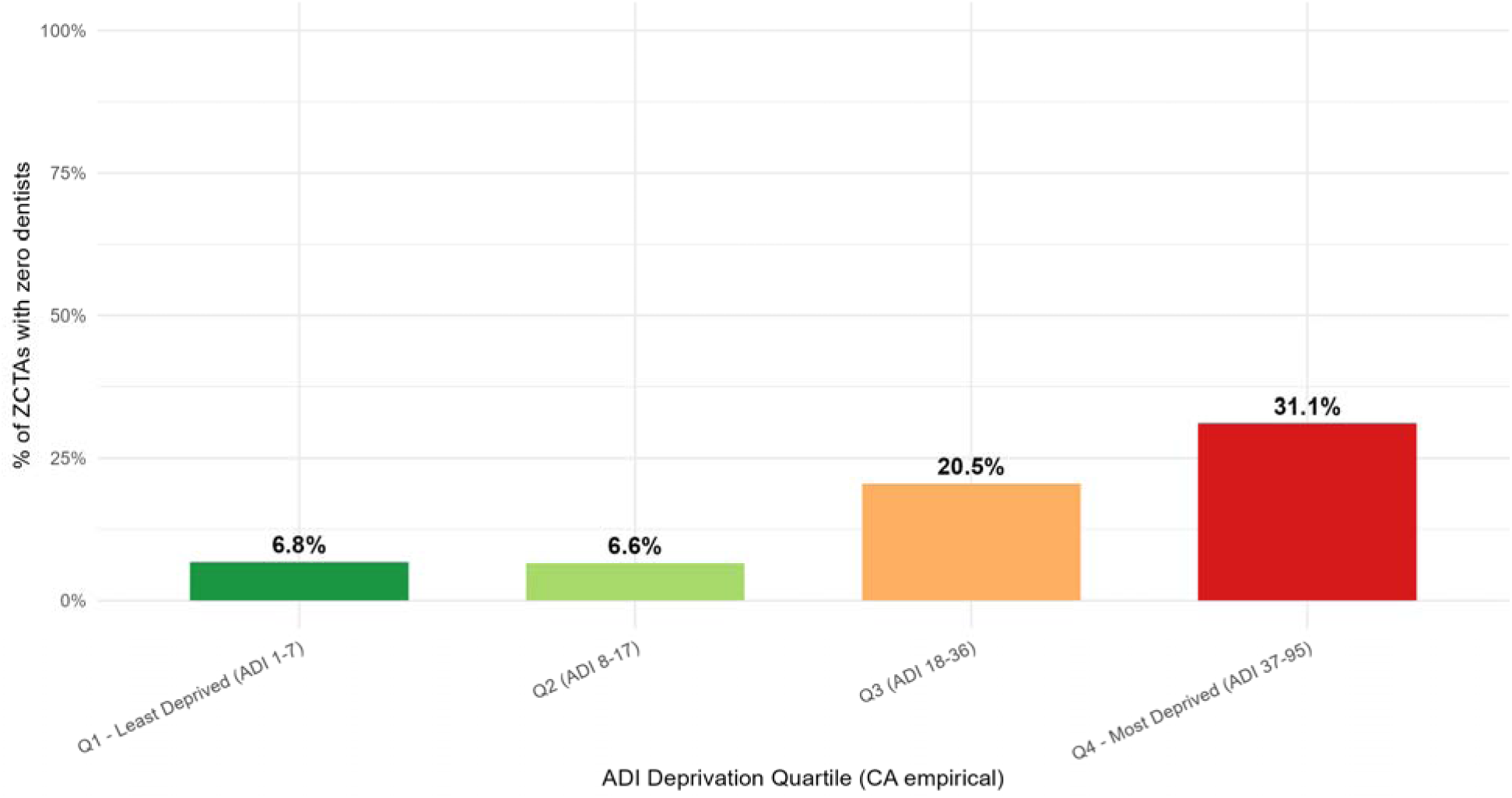
Percentage of California ZCTAs with zero active dental providers, by California-specific empirical ADI quartile.

### Specialist vs. General Dentist Availability

Across the analytic sample, 31,868 general dentists (84.0%) and 6,077 dental specialists (16.0%) were identified. Both provider groups exhibited monotonic gradients across California-specific ADI quartiles. Mean general-dentist density declined from 11.56 per 10,000 residents in the least-deprived quartile (Q1) to 5.08 per 10,000 in the most-deprived quartile (Q4), representing a 56% reduction. Mean specialist density declined from 2.92 to 0.42 per 10,000 residents across the same quartiles, representing an 86% reduction. The prevalence of ZCTAs without any dental specialist was consistently higher than the prevalence of ZCTAs without a general dentist across all deprivation levels. In the most-deprived quartile, 73.9% of ZCTAs contained no dental specialist compared with 31.4% containing no general dentist. The average specialist share of the local dental workforce declined from 16.9% in Q1 to 6.1% in Q4. Overall, 42.5% of ZCTAs contained no dental specialist, compared with 16.2% that contained no general dentist. These findings suggest that geographic disparities in dental workforce availability extend to both general and specialty care and may be particularly pronounced for specialist services (Supplementary Table S2).

### Robustness and Sensitivity Analyses

The negative association was robust to the choice of model family. A log-linear ordinary least squares regression of log(dentists per 10,000 + 0.1) on the median ADI national rank produced a coefficient of −0.0314 (95% CI −0.0356 to −0.0272; p < 0.001), corresponding to a multiplicative decrease in density of approximately 3.1% per ADI percentile on the log scale. The coefficient was numerically stable across alternative zero-handling constants (0.01, 0.1, and 1.0).

Excluding Travis Air Force Base (ZIP 94535), the single military installation in the analytic sample with embedded dental services, did not materially change the primary estimate (RR 0.9823; 95% CI 0.9799 to 0.9847; p < 0.001), indicating that the deprivation gradient was not driven by this outlier. Replacing the continuous national ADI percentile with the California-specific state decile (1–10, where 10 indicates greatest within-state deprivation) yielded a RR of 0.8822 per decile (95% CI 0.8679 to 0.8967; p < 0.001), consistent with the direction and magnitude of the primary finding. Across all specifications, the deprivation gradient remained negative, statistically significant, and of comparable magnitude.

### County-Level Geographic Patterns

ZCTA-level results were aggregated to the California county level for visualization using the U.S. Census Bureau 2020 ZCTA-to-county relationship file (Figure 6). The two side-by-side choropleth maps showed a spatial correspondence between county-level provider density and county-level deprivation. Counties with the highest dental provider densities were concentrated in the lower-deprivation San Francisco Bay Area and surrounding coastal counties, including San Francisco (16.0 per 10,000; ADI 2.7), Santa Clara (13.4 per 10,000; ADI 3.8), Marin, San Mateo, and Orange. Counties with the lowest densities were located in the higher-deprivation rural northern and far-eastern interior of the state, including Trinity (1.1 per 10,000; ADI 49), Modoc (3.7; ADI 84), Imperial (4.1; ADI 60), and Inyo (1.5; ADI 58). The spatial pattern was consistent with the negative association observed in the ZCTA-level analyses.

**Figure 6.**
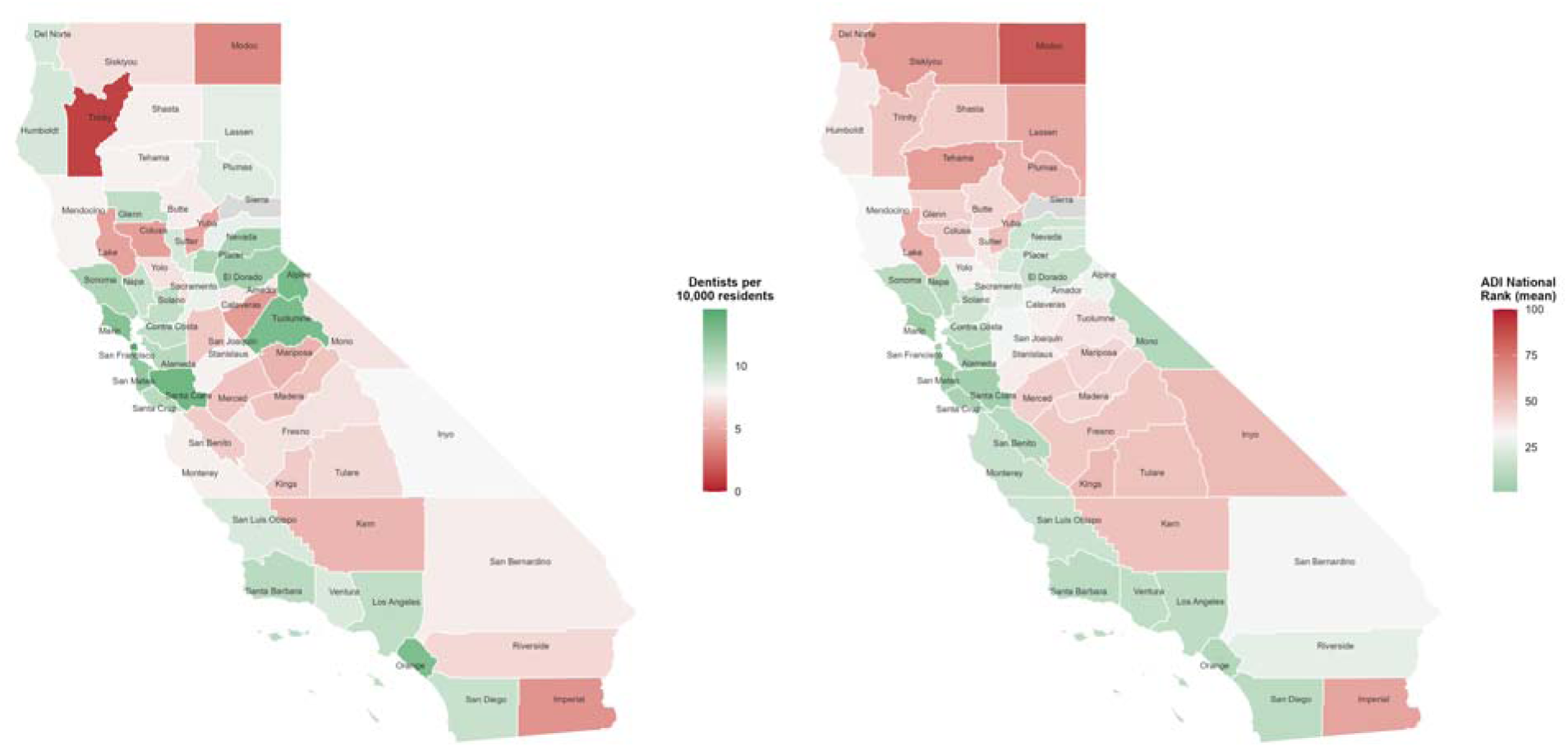
Dental provider density (left) and area deprivation (right) by California county. *Note: Both maps use a shared red–white–green diverging palette anchored at the California county median, with red indicating the less favorable value on each map (fewer dentists on the left; greater deprivation on the right) and green indicating the more favorable value. Counties near the state median appear near-white on both maps. Sierra County is shown in grey owing to insufficient analytic data. ZCTA-level data (population* ≥ *500) were aggregated to counties using the Census ZCTA-to-county crosswalk; dentist density was winsorized at the 99th percentile (14.5 per 10,000) for the color scale*.

## Discussion

In this statewide ecological analysis of California, greater neighborhood deprivation was consistently associated with lower dental provider density, even within a state that has the largest dental workforce in the United States. These findings suggest that overall dental workforce supply alone is an incomplete indicator of access to care. Instead, where providers practice relative to neighborhood socioeconomic conditions appears to play a critical role in determining access. Communities with greater deprivation not only had substantially fewer dental providers but were also more likely to be dental deserts, indicating that socioeconomic disadvantage and workforce shortages frequently coexist. Together, these findings suggest that strategies focused solely on increasing the overall number of dentists may be insufficient unless they also address the geographic distribution of the dental workforce.

A key contribution of this study is its use of ZCTAs as the unit of analysis. Compared with county-level analyses, this approach provides greater geographic granularity and allows identification of neighborhood-level disparities in dental provider availability that may otherwise be obscured within larger administrative units. This is particularly important in California, where substantial socioeconomic heterogeneity can exist within the same county. By identifying disparities at the neighborhood level, these findings may better inform targeted workforce planning and resource allocation.

Our findings are consistent with previous studies demonstrating geographic inequities in dental care access. Rahman et al. identified dental clinic deserts across the United States, Benavidez et al. reported concordance between dental shortage area designations and social vulnerability, and Gaskin et al. linked greater provider availability with lower emergency dental care utilization (8,15). However, most previous studies have relied on county-level analyses, broader measures of social vulnerability, or measures of realized access rather than provider supply (14). By examining dental provider density in relation to the Area Deprivation Index at the ZCTA level, the present study captures neighborhood-scale differences that are often masked by larger geographic units. For example, although Los Angeles County has an overall dental provider density above federal shortage thresholds (25), Beverly Hills ZCTAs in the analytic sample contained 71–132 active dental providers per 10,000 residents, whereas South Los Angeles ZCTAs less than 13 miles away, including ZIP 90011 (population >100,000), contained fewer than three providers per 10,000 residents. These intra-county disparities are largely invisible in county-level analyses and illustrate how county-level shortage designations may overlook neighborhoods with substantial unmet workforce needs within otherwise well-supplied jurisdictions. This neighborhood-level perspective may therefore help identify communities where workforce shortages intersect with socioeconomic disadvantage and improve the targeting of workforce planning and resource allocation.

The findings are particularly relevant in California, which has one of the largest dental workforces in the United States and the nation’s largest Medicaid dental program. Despite these substantial workforce and coverage resources, marked disparities in provider availability persisted across communities. The coexistence of a large statewide dental workforce with persistent neighborhood-level shortages highlights the importance of where providers practice, not simply how many providers are available statewide. Policies focused solely on increasing the number of dental providers may not fully address disparities if providers continue to cluster in more advantaged communities. Workforce distribution may therefore be as important as workforce supply in determining access to care. Uneven provider distribution may also contribute to broader oral health disparities by limiting access to preventive services, delaying treatment, and increasing reliance on urgent or emergency dental care in disadvantaged communities.

A particularly noteworthy finding was that disparities were considerably larger for dental specialists than for general dentists. While general dentist density declined by 56% across deprivation quartiles, specialist density declined by 86%. Nearly three-quarters of the most deprived ZCTAs lacked any dental specialist. These findings suggest that socioeconomic disparities in workforce distribution may disproportionately limit access to advanced dental services such as endodontics, pediatric dentistry, oral surgery, and periodontics. Reduced specialist availability may contribute to delayed treatment, increased travel burden, and reduced access to complex dental care. The substantially steeper deprivation gradient observed for specialists suggests that workforce inequities may be even more pronounced for specialty services than for general dental care.

The findings suggest that the Area Deprivation Index may serve as a useful planning tool for identifying communities at elevated risk of dental workforce shortages. Because ADI incorporates multiple dimensions of socioeconomic disadvantage, it may provide a more comprehensive measure of community need than provider-to-population ratios alone. Policymakers could use ADI-informed approaches to prioritize workforce incentives, loan repayment programs, residency rotations, and community-based practice initiatives in underserved areas. Such approaches may help direct limited workforce resources toward communities with the greatest need and reduce persistent geographic disparities in access to dental care.

Future research should examine whether similar deprivation gradients exist for specific dental specialties beyond the descriptive sub-analysis presented here, Medicaid-participating providers, and measures of realized access. More sophisticated spatial accessibility approaches, such as enhanced two-step floating catchment area methods applied nationally by Rahman et al. and county-level drive-time analyses similar to those used by the California Health Care Foundation for maternity services, could further characterize how the deprivation gradient identified in this study translates into travel burden and realized access to dental care. Longitudinal analyses may also help determine whether changes in neighborhood deprivation are associated with subsequent changes in dental workforce distribution and whether these changes reduce geographic disparities in access over time.

### Strengths and Limitations

This study has several strengths. By examining neighborhood deprivation in relation to dental provider density using the Area Deprivation Index (ADI) and National Plan and Provider Enumeration System (NPPES) data, the study provides a contemporary statewide assessment of geographic disparities in California. The analysis included 37,945 active dental providers across 1,426 ZIP Code Tabulation Areas (ZCTAs), representing approximately 39 million California residents. The use of publicly available national datasets enhances the transparency, reproducibility and comparability of the study. In addition, the observed deprivation gradient remained consistent across multiple model specifications, continuous and categorical ADI analyses, and prespecified sensitivity analyses, supporting the robustness of the findings.

Despite the statewide scope and comprehensive provider coverage of this analysis, several methodological considerations should be acknowledged. NPPES records provider practice locations rather than patient residences; therefore, provider density may not fully reflect residential access to care in some areas. Because provider locations are reported by ZIP code and population estimates are available at the ZCTA level, ADI values were aggregated from the Neighborhood Atlas ZIP+4 crosswalk to ZCTAs, which may mask within-area socioeconomic variation. However, any resulting exposure misclassification would likely be non-differential and bias associations toward the null. NPPES also reflects provider registration rather than actual service utilization, appointment availability, insurance acceptance, or patient travel patterns and may include providers who are retired or practicing only part-time. Accordingly, provider density should be interpreted as a measure of workforce availability rather than realized access to care.

## Conclusion

In conclusion, neighborhood deprivation was consistently associated with lower dental provider density across California. Communities experiencing greater socioeconomic disadvantage had substantially fewer dental providers, were more likely to be dental deserts, and experienced particularly pronounced shortages of dental specialists. These findings suggest that significant disparities in workforce availability can persist even within a state that has one of the largest dental workforces in the nation, highlighting the importance of workforce distribution in addition to overall workforce supply. The findings further suggest that the Area Deprivation Index may be a useful tool for identifying communities where dental workforce shortages intersect with broader socioeconomic disadvantage. ADI-informed workforce planning strategies may help promote a more equitable distribution of the dental workforce and reduce geographic disparities in access to oral health care.

## Data Availability

All datasets analyzed during this study are publicly available. Dental provider data were obtained from the National Plan and Provider Enumeration System (NPPES) maintained by the Centers for Medicare & Medicaid Services. Area Deprivation Index data were obtained from the Neighborhood Atlas. Population estimates were obtained from the U.S. Census Bureau. The analytic code is available from the corresponding author upon reasonable request.

https://download.cms.gov/nppes/NPI_Files.html

https://www.neighborhoodatlas.medicine.wisc.edu/

https://www.census.gov/

## Abbreviations

ADI: Area Deprivation Index
CI: Confidence Interval
CMS: Centers for Medicare & Medicaid Services
RR: Rate Ratio
NPI: National Provider Identifier
NPPES: National Plan and Provider Enumeration System
STROBE: Strengthening the Reporting of Observational Studies in Epidemiology
ZCTA: ZIP Code Tabulation Area

## Declarations

### Ethics approval and consent to participate

This study used publicly available, de-identified, and aggregated data from the National Plan and Provider Enumeration System (NPPES), the Neighborhood Atlas, and the United States Census Bureau. No human participants were involved, and no identifiable individual-level data were collected or analyzed. Therefore, this study did not constitute human subjects research and institutional review board approval and informed consent were not required.

### Consent for publication

Not applicable.

### Availability of data and materials

All datasets analyzed during the current study are publicly available. Dental provider data were obtained from the National Plan and Provider Enumeration System (NPPES) database maintained by the Centers for Medicare & Medicaid Services. Area Deprivation Index data were obtained from the Neighborhood Atlas. Population estimates were obtained from the United States Census Bureau. The analytic code is available from the corresponding author upon reasonable request.

### Authors’ contributions

ALA conceived the study, conducted the literature review, interpreted findings, and drafted the manuscript. CG contributed to study conception, methodology, data analysis, visualization, and manuscript revision. Both authors read and approved the final manuscript.

## Acknowledgements

The authors thank Dr. Benjamin W. Chaffee for his valuable feedback and review of the manuscript.

---

**Supplementary Table S1.** California dental providers by National Provider Identifier (NPI) taxonomy code, final analytic sample (n = 37,945).

| **Taxonomy code** | **Taxonomy label** | **N providers** | **% of total** |
| --- | --- | --- | --- |
| **General Dentistry** |  |  |  |
| 1223G0001X | General Practice | 17,404 | 45.9 |
| 122300000X | General Dentist | 14,464 | 38.1 |
| **Dental Specialties** |  |  |  |
| 1223X0400X | Orthodontics and Dentofacial Orthopedics | 1,724 | 4.5 |
| 1223P0221X | Pediatric Dentistry | 1,163 | 3.1 |
| 1223E0200X | Endodontics | 890 | 2.3 |
| 1223S0112X | Oral and Maxillofacial Surgery | 861 | 2.3 |
| 1223P0300X | Periodontics | 809 | 2.1 |
| 1223P0700X | Prosthodontics | 381 | 1.0 |
| 1223D0001X | Dental Public Health | 104 | 0.3 |
| 1223P0106X | Oral and Maxillofacial Pathology | 64 | 0.2 |
| 1223D0004X | Dental Anesthesiology | 54 | 0.1 |
| 1223X2210X | Orofacial Pain | 15 | <0.1 |
| 1223X0008X | Oral and Maxillofacial Radiology | 11 | <0.1 |
| 125Q00000X | Oral Medicine | 1 | <0.1 |
|  | **Total dental providers** | **37,945** | **100.0** |

Notes: Counts reflect providers in the final analytic sample (n = 37,945 across 1,426 ZCTAs). The pre-exclusion NPPES extract contained 39,049 providers; 1,104 (2.8%) were dropped under the ZCTA-level exclusion criteria described in the Methods. Each provider was assigned a single primary dental taxonomy code. NPPES = National Plan and Provider Enumeration System.

**Supplementary Table S2.** Descriptive comparison of general dentists and dental specialists across California ZCTAs, overall and by California-specific empirical ADI deprivation quartile.

| **ADI quartile** | **N ZCTAs** | **General dentists** |  |  | **Dental specialists** |  |  | **Specialist share (%)** |
| --- | --- | --- | --- | --- | --- | --- | --- | --- |
|  |  | **N** | **Mean / 10k** | **% with none** | **N** | **Mean / 10k** | **% with none** |  |
| Q1 (ADI 1–7) | 384 | 11,693 | 11.56 | 7.0 | 2,847 | 2.92 | 20.1 | 16.9 |
| Q2 (ADI 8–17) | 363 | 10,560 | 8.93 | 6.6 | 1,903 | 1.58 | 26.2 | 12.2 |
| Q3 (ADI 18–36) | 322 | 6,229 | 6.21 | 21.1 | 957 | 0.98 | 52.8 | 9.3 |
| Q4 (ADI 37–95) | 357 | 3,386 | 5.08 | 31.4 | 370 | 0.42 | 73.9 | 6.1 |
| **Overall (all ZCTAs)** | **1,426** | **31,868** | **8.06** | **16.2** | **6,077** | **1.52** | **42.5** | **16.0** |
| **Q1 → Q4 change** | — | — | –56% | +24.4 pp | — | –86% | +53.8 pp | –10.8 pp |

Notes: Densities are providers per 10,000 residents, computed at the ZCTA level and averaged across ZCTAs within each group. “N” columns show the total number of providers of each type. “% with none” indicates the percentage of ZCTAs in each group containing no provider of the indicated type. Specialist share denotes the average percentage of the local dental workforce
*composed of specialists, calculated within ZCTAs containing at least one dental provider. Contrasts in the bottom row express the absolute (percentage point, pp) or relative (%) change from Q1 to Q4. ADI = Area Deprivation Index.*

